# Towards automated multi-regional lung parcellation for 0.55-3T 3D T2w fetal MRI

**DOI:** 10.1101/2025.06.22.25330066

**Authors:** Alena U. Uus, Carla Avena Zampieri, Fenella Downes, Alexia Egloff Collado, Megan Hall, Joseph Davidson, Kelly Payette, Jordina Aviles Verdera, Irina Grigorescu, Joseph V. Hajnal, Maria Deprez, Michael Aertsen, Jana Hutter, Mary A. Rutherford, Jan Deprest, Lisa Story

**Affiliations:** Biomedical Engineering Department, King’s College London, St. Thomas’ Hospital, London, United Kingdom; Centre for the Developing Brain, King’s College London, London, UK; Department of Women and Children’s Health, King’s College London, London, UK; Department of Paediatric Surgery, Evelina Children’s Hospital, London, UK; GOS-UCL Institute of Child Health, London,UK; Department of Imaging and Pathology, Clinical Department of Radiology, University Hospitals KU Leuven, Leuven, Belgium; Smart Imaging Lab, Radiological Institute, University Hospital Erlangen, Germany; Clinical Department of Obstetrics and Gynaecology, University Hospitals Leuven, Leuven, Belgium; Elizabeth Garrett Anderson Institute of Women’s Health, University College London, London, UK; Fetal Medicine Unit, Guy’s and St Thomas’ NHS Foundation Trust, London, UK

**Keywords:** Fetal MRI, Lung segmentation, Normal lung development, Congenital diaphragmatic hernia

## Abstract

Fetal MRI is increasingly being employed in the diagnosis of fetal lung anomalies and segmentation-derived total fetal lung volumes are used as one of the parameters for prediction of neonatal outcomes. However, in clinical practice, segmentation is performed manually in 2D motion-corrupted stacks with thick slices which is time consuming and can lead to variations in estimated volumes. Furthermore, there is a known lack of consensus regarding a universal lung parcellation protocol and expected normal total lung volume formulas. The lungs are also segmented as one label without parcellation into lobes. In terms of automation, to the best of our knowledge, there have been no reported works on multi-lobe segmentation for fetal lung MRI. This work introduces the first automated deep learning segmentation pipeline for multi-regional lung segmentation for 3D motion-corrected T2w fetal body images for normal anatomy and congenital diaphragmatic hernia cases. The protocol for parcellation into 5 standard lobes was defined in the population-averaged 3D atlas. It was then used to generate a multi-label training dataset including 104 normal anatomy controls and 45 congenital diaphragmatic hernia cases from 0.55T, 1.5T and 3T acquisition protocols. The performance of 3D Attention UNet network was evaluated on 18 cases and showed good results for normal lung anatomy with expectedly lower Dice values for the ipsilateral lung. In addition, we also produced normal lung volumetry growth charts from 290 0.55T and 3T controls. This is the first step towards automated multi-regional fetal lung analysis for 3D fetal MRI.

## 1 Introduction

Fetal lung development is a complex process that is crucial in determining neonatal outcomes, especially in cases involving hypoplasia-related anomalies like congenital diaphragmatic hernia (CDH) [26]. Normal lung development involves intricate development of the bronchial tree and increase in size and differentiation into lobes, critical for efficient respiratory function postnatally [8]. This complex process encompasses a series of orchestrated stages during which lung lobes form through branching morphogenesis, where lung buds outpouch from the foregut endoderm, elongate, and branch into distinct lobes [18]. Imaging modalities have become invaluable tools in evaluating fetal lung development. Fetal MRI, in addition to prenatal ultrasound, has been increasingly employed for diagnosis of fetal lung anomalies [6,2] as well as characterisation of development patterns [11,19]. In T2w MR images [1] fetal lung tissue appears moderately hyperintense due to its high fluid content and has the optimal contrast with surrounding organs for delineation of lungs to extract volumetric information. e.g., segmentation-derived total lung volume (TLV) is used in clinical practice for quantification of observed to expected TLV in CDH cases [9]. Parcellation of the lungs is generally performed manually slice-by-slice in several acquisition planes (e.g., axial) using classical vendor-specific tools [15]. Segmentation time and reproducibility depend on the acquisition parameters, fetal size and level of expertise.

While modern sequences allow fast acquisition of individual slices, raw MRI stacks are frequently corrupted by both fetal and maternal motion that leads to loss of structural continuity in 3D that naturally affects quality of segmentation and the reliability of volumetry results. This can be resolved by retrospective motion correction methods [22] based on deformable slice-to-volume registration (DSVR) for image-domain reconstruction of 3D images from multiple motion-corrupted stacks. In addition, the 3D DSVR images are reoriented in the standard radiological space that makes both segmentation and visual analysis significantly easier [21]. [7] showed that 3D DSVR-derived segmentations produce comparable lung volumetry to 2D slice-wise segmentations.

In terms of consistent application of fetal MRI lung volumetry in clinical practice, there is a reported lack of consensus on formalisation of lung segmentation protocols and techniques between different clinical centres [14] (e.g., which acquisition plane should be used, exclusion of vessels, impact of motion-corruption, etc.). A recent review also reported that using different expected TLV (eTLV) model formulas might affect predicted outcomes and counselling for CDH [27]. Furthermore, the existing eTLV formulas were all created based on segmentation of clinical fetal MRI datasets without lung anomalies but with other anomalies rather than true normal controls. Recently, [13,23] confirmed the feasibility of deep learning for automated segmentation of the lungs in 3D motion-corrected T2w and T2* fetal MRI body images. The left and right lungs were segmented as one combined label and only normal anatomy cases from the same acquisition protocols were used. Conte et al. [5] successfully applied 2D nnUNet for left/right lung segmentation in raw T2w MRI stacks. Yet, they reported suboptimal results for the ipsilateral lung potentially due to motion and thick slices. This highlights the need for both formalisation of a universal standard fetal lung parcellation protocol as well as development of tools for robust automated multi-regional segmentation for more comprehensive analysis.

### Contributions

In this work, we propose the first solution for automated multi-lobe lung volumetry of 3D motion-corrected structural T2w fetal body MRI for both normal and abnormal lung anatomy. We build upon the existing 3D T2w structural body organ segmentation pipeline [23] by formalising a novel multi-lobe lung parcellation protocol and training a deep learning segmentation network for 0.55-3T normal lung anatomy. We also added 45 multi-centre CDH datasets to the training in order to assess the general feasibility of 3D automated segmentation for abnormal anatomy. The final trained network was used to generate growth charts of the normal lung development from 290 control subject datasets at 0.55T and 3T field strengths.

## 2 Methods

### 2.1 Cohort, acquisition and pre-processing

This study employed fetal MRI data acquired at two different sites: at St.Thomas’ Hospital, London, UK under the “Individualised risk prediction of adverse neonatal outcome in pregnancies that deliver preterm using advanced MRI techniques and machine learning” study (REC 21/SS/0082), “NANO” (REC 22/YH/0210), “MEERKAT” (REC 21/LO/0742), “Placenta Imaging Project” (REC 16/LO/1573), “Quantification of fetal brain growth and development using MRI” (REC 07/H0707/105), “IFIND” (REC 14/LO/1806) studies and at University Hospitals Leuven, Belgium (Ethics number: S5678). It includes 306 normal lung anatomy datasets from 16 to 39 weeks GA range and 51 datasets of subjects with CDH from 22 to 33 weeks GA range.

The T2w datasets were acquired on 5 different scanners: 0.55T Siemens Free.Max MRI, TE=105ms, 1.48×1.48mm resolution, 4.5mm slice thickness; 1.5T Philips Ingenia MRI, TE=80/180ms, 1.25×1.25mm resolution, 2.5mm slice thickness, −1.25mm gap; 1.5T Siemens Sola MRI, TE=80ms, 1.25×1.25mm resolution, 3.0mm slice thickness; 1.5T Aera Siemens MRI, TE=90ms, 0.74×0.74mm resolution, 3.0-4.00mm slice thickness; 3T Philips Achieva MRI, TE=180ms, 1.25×1.25mm resolution, 2.5mm slice thickness, −1.5mm gap. For all datasets, we used the SVRTK docker ^11^ with either fully automated [21] or manual [20] DSVR method to reconstruct 3D isotropic images of the fetal body from multiple motion-corrupted T2w stacks (4-8 per case). The output images have 0.75-0.8 mm isotropic resolution and are reoriented in the standard space[23]. The main selection criteria were good reconstruction image quality and a singleton pregnancy. Taking into account the limited number of CDH cases, we used all available cases including left and right CDH cases with different degree of severity and presence of additional various lesions. The CDH datasets were acquired predominantly on 1.5T.

### 2.2 Formalisation of multi-regional lung parcellation protocol

The fetal multi-regional parcellation protocol was formalised based on the standard lung anatomy [18] and includes 5 lobes: superior left, inferior left, superior right, middle right and inferior right. In addition, we also included extra labels for other internal thorax structures in order to improve the accuracy of lung segmentation: thymus, heart and major vessels, trachea, esophagus and hyper-intense lesions (10 labels in total). The parcellation was performed manually in the 3D T2w averaged fetal body atlas space [23] using ITK-SNAP^12^ by 3 clinicians and a researcher with extensive experience in fetal MRI. Since lung lobe fissures are not normally visible on fetal MRI images, the parcellation was guided by the location of bronchi and pulmonary vessel branches [10] and regions with no pronounced vasculature, fetal lung histology studies [24] and annotated adult CT lung lobe segmentation datasets [25]. The parcellation label file is be publicly available online at the 3D fetal body MRI atlas repository ^13^.

### 2.3 Automated multi-regional lung segmentation

We used the standard MONAI [4] 3D Attention-UNet [12] implementations with five and four encoder-decoder blocks (output channels 16, 32, 64, 128, 256), convolution and upsampling kernel size of 3, ReLU activation, dropout ratio of 0.5. We employed AdamW optimiser with a linearly decaying learning rate, initialised at 1 × 10^−3^, default *β* parameters and weight decay=1 × 10^−5^. Augmentation included the standard MONAI affine rotations, contrast adjustment and bias field and cropping to the thorax ROI. Preprocessing included rescaling to 0-1 range and resampling with padding to 256×256×256 voxel grid. The training dataset was created in two steps. At first, the atlas labels were propagated to a preliminary small set of 20 0.55T/3T normal and 10 CDH 3D images using affine and non-rigid (local cross-correlation similarity metric; default settings) MIRTK package ^14^. This was followed by manual refinement in ITK-SNAP. For CDH cases, the ipsilateral lung was segmented as the superior lobe in the corresponding affected side and all hyperintense lesions (e.g., pulmonary airway malformation) were segmented as an extra label. These labels were then used to train a preliminary version of the network with 5000 iterations. Next, we used the preliminary network weights to segment the second version of the training dataset with 104 normal anatomy and 45 CDH cases that were inspected and manually refined. The final training was performed with 10000 iterations. The testing of the network performance was tested on 12 0.55T/3T normal control and 6 CDH cases (mixture of 0.55T/1.5T/3T protocols)from different GA ranges vs. manual labels. The trained network weights and segmentation script are publicly available online as a standalone docker application in repository^15^.

### 2.4 Normal lung volumetry growth charts

Next, we used the network to segment 135 0.55T and 155 3T normal control datasets from 16 - 39 weeks GA range. The inclusion criteria were no reported fetal, placental or maternal anomalies. All segmentations were reviewed and manually refined, if required. The extracted lobe volumes were used to generate normative growth charts with mean, 5th and 95th centiles [16] and assess the global volumetry trends. The impact of acquisition parameters was analysed using ANCOVA from Python statsmodels module.

## 3 Experiments and Results

### 3.1 Proposed multi-regional lung parcellation protocol

The proposed lung parcellation protocol is shown in Fig. 1 (the network output for the 3T population-averaged fetal atlas). The parcellation map was inspected by clinicians with extensive experience in fetal MRI and confirmed to be acceptable. The label boundaries correspond to the expected position of the lobes with respect to the main branches of bronchi and pulmonary vasculature. There are no pronounced vascular structures crossing the interfaces between the lobes used as an indirect indication of the location of the fissures. However, in this case, we could not evaluate the impact of the occurrence of normal variants like azygos fissure, left horizontal fissure, etc. which will be the subject for future work.

**Fig. 1.**
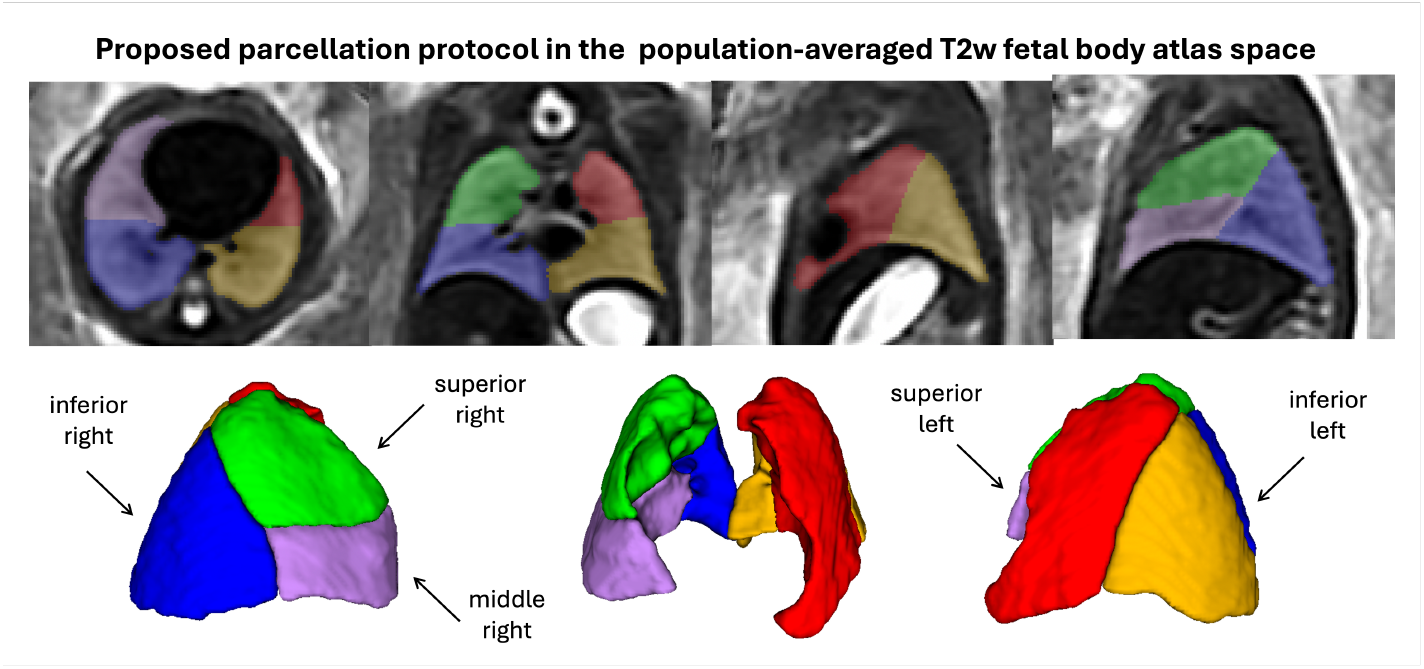
Proposed 3D multi lobe lung parcellation protocol in the T2w population-averaged fetal body atlas space.

### 3.2 Automated multi-regional lung segmentation

Tab. 1 summarises the results of testing of automated segmentation on 12 control and 6 CDH (3 right and 3 left) cases from 0.55/1.5/3T acquisition protocols and 22 - 36 weeks GA range. The examples of auto segmentation results are shown in Fig. 2. Quantitative evaluation was performed vs. manual labels for individual and combined lobes. The network outputs have relatively adequate Dice values for all lobes for the normal control cases with better performance for the 3T cohort due to lower visibility of features, blurring and lower SNR in 0.55T cases. Visual inspection of the results confirmed that the lobes were segmented appropriately, however the network performed better for cases with higher lung tissue contrast (later GA, 3T). This confirms the general feasiblity of multi-regional 3D lung segmentation for different acquisition protocols.

**Table 1.** Testing of the segmentation network on 12 control (0.55T and 3T), 3 right CDH and 3 left CDH cases (mixture of 0.55T, 1.5T and 3T protocols): Dice values per ROI vs. manual labels. The values corresponding to ipsilateral lungs are highlighted in blue.

| ROI | 3T controls | 0.55T controls | Left CDH | Right CDH |
| --- | --- | --- | --- | --- |
| Both lungs combined | $0.920 \pm 0.019$ | $0.915 \pm 0.018$ | $0.873 \pm 0.032$ | $0.820 \pm 0.083$ |
| Left lung combined | $0.919 \pm 0.023$ | $0.899 \pm 0.020$ | $0.709 \pm 0.045$ | $0.846 \pm 0.079$ |
| Right lung combined | $0.919 \pm 0.018$ | $0.921 \pm 0.025$ | $0.896 \pm 0.032$ | $0.572 \pm 0.056$ |
| Left superior lobe | $0.868 \pm 0.028$ | $0.839 \pm 0.051$ | $0.709 \pm 0.045$ | $0.793 \pm 0.065$ |
| Left inferior lobe | $0.914 \pm 0.021$ | $0.887 \pm 0.027$ | - | $0.838 \pm 0.079$ |
| Right superior lobe | $0.875 \pm 0.021$ | $0.867 \pm 0.031$ | $0.840 \pm 0.049$ | $0.572 \pm 0.056$ |
| Right middle lobe | $0.852 \pm 0.009$ | $0.843 \pm 0.032$ | $0.734 \pm 0.056$ | - |
| Right inferior lobe | $0.906 \pm 0.023$ | $0.908 \pm 0.029$ | $0.879 \pm 0.015$ | - |

**Fig. 2.**
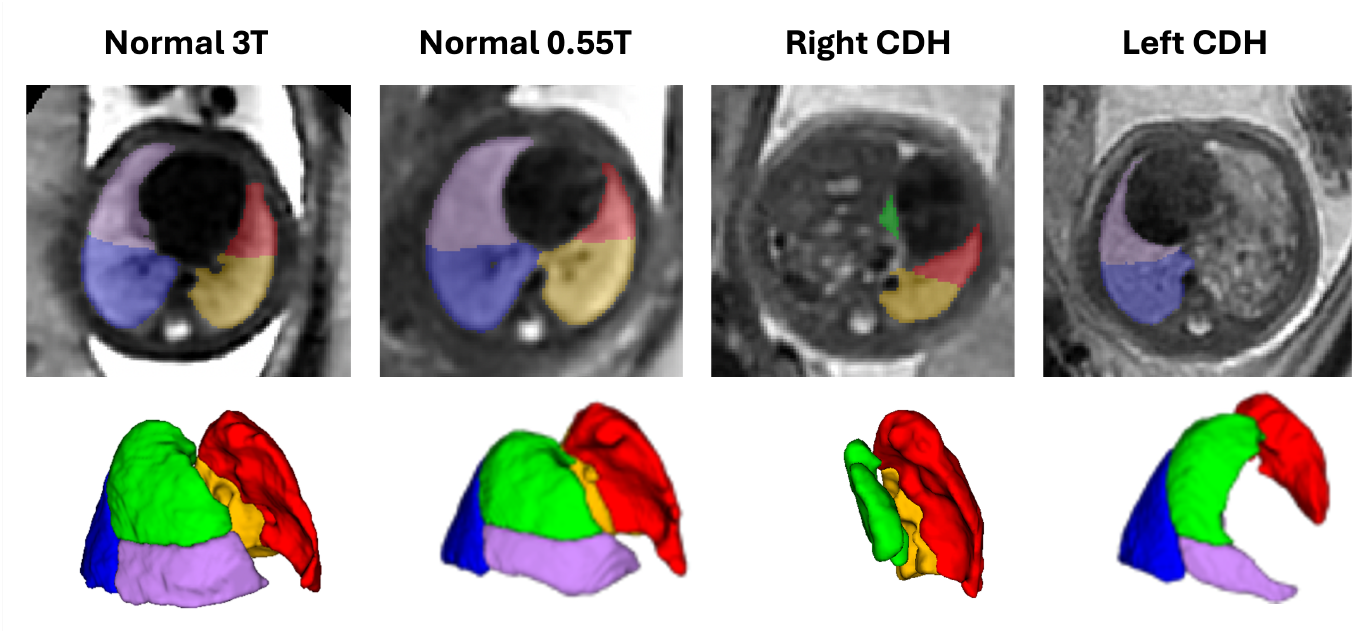
Examples of automated segmentation results for 3T and 0.55T normal controls and 1.5T left and right CDH cases.

The expected lower Dice range for the ipsilateral lung and the CDH cases emphasises the need for more extensive training datasets with a larger number and diversity of abnormal anatomy, which, significantly varies from case to case in CDH. In particular, the poorer results for right CDH can also be explained as the image quality of the available right CDH test cases was suboptimal with limited visibility and contrast and imperfections in manual segmentations. Further development of this pipeline would require more precise definition of the abnormal features and a more advanced deep learning approach as well as extension of the parcellation protocol with a dedicated ipsilateral label.

### 3.3 Normal lung volumetry growth charts

The combined total lung volumes graphs (both 0.55T and 3T) in Fig. 3.A show relatively good correlation with the previously proposed models based on manual 2D slice labels. The higher degree of variance at later GA is also in accordance with the previously reported works [17,3,11]. Unlike previous works, this study included only normal control fetuses without other reported fetal or maternal anomalies further stressing the need for a more complex and patient-specific expected TLV estimation approach.

**Fig. 3.**
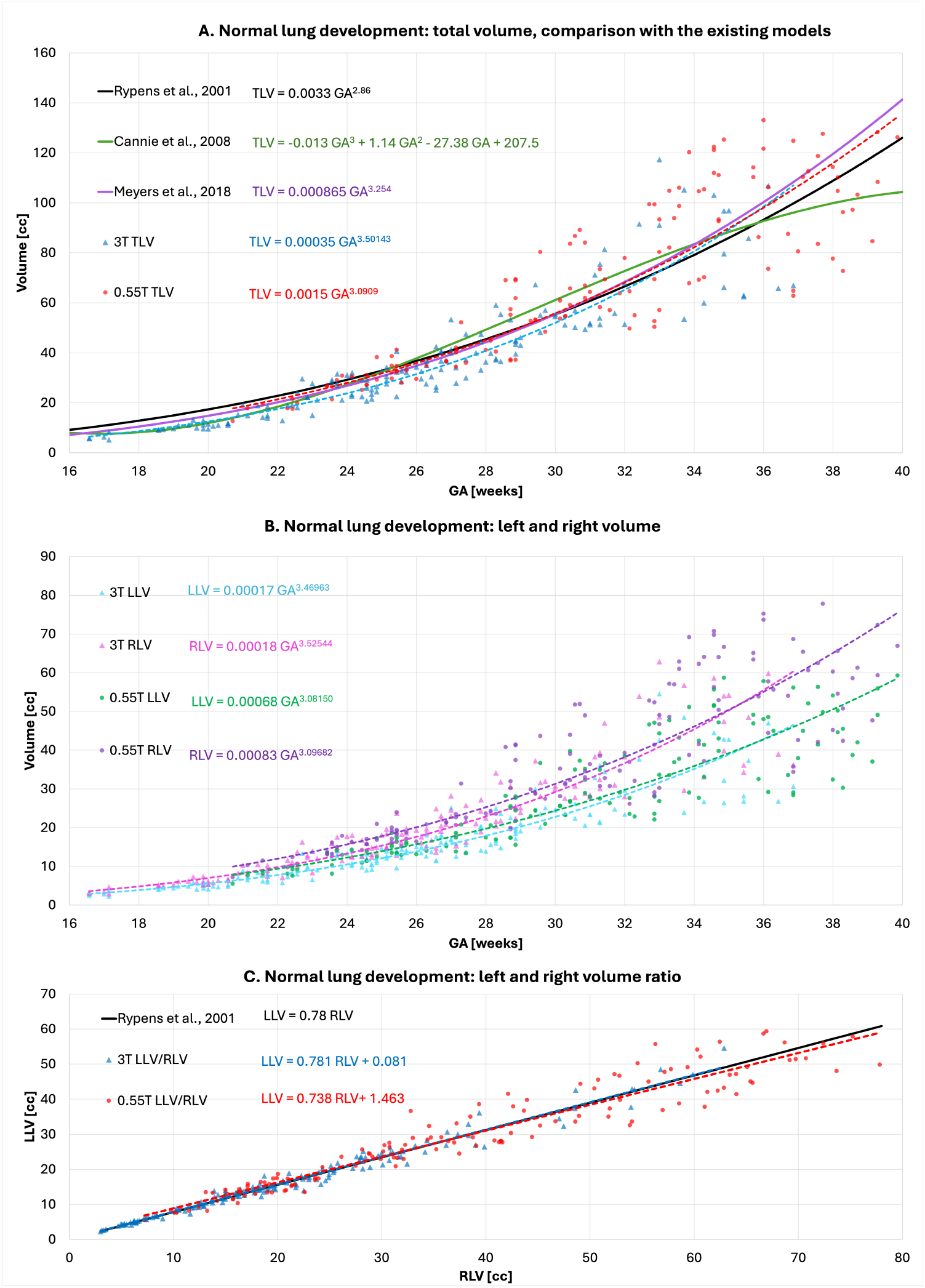
Growth charts created from 155 3T and 135 0.55T normal fetal control 3D T2w datasets for 16 to 39 week GA range: total lung volume (TLV) (A); left (LLV) and right (RLV) lung volumes (B); left and right volume ratio (C).

There is a significant difference (p<0.001) between the 0.55T and 3T TLV. Smaller 3T TLV values vs. 0.55T could be potentially related to the large difference in the echo time that has a visible effect on the contrast interface between the lung and surrounding tissue, exclusion of vessels as well as the partial volume impact due to the larger voxel size in 0.55T datasets. This suggests that any studies of abnormal datasets with mixed acquisition parameters require a matched mixture of acquisition parameters in control datasets. The left and right lung volumes also show the increasing variance with GA in both 0.55T and 3T (Fig. 3.B) and slightly lower values for the 3T cohort. The ratio between left to right lung volume is also similar to the previously reported by Rypens et al. [17]. The graphs in Fig. 4 demonstrate the changes in lung lobe volumes vs. GA for normal control 0.55T and 3T datasets with expected increase in volumes and stable relative proportions between the lobes. The document with centile formulas is publicly available online at the fetal body MRI atlas repository^16^.

**Fig. 4.**
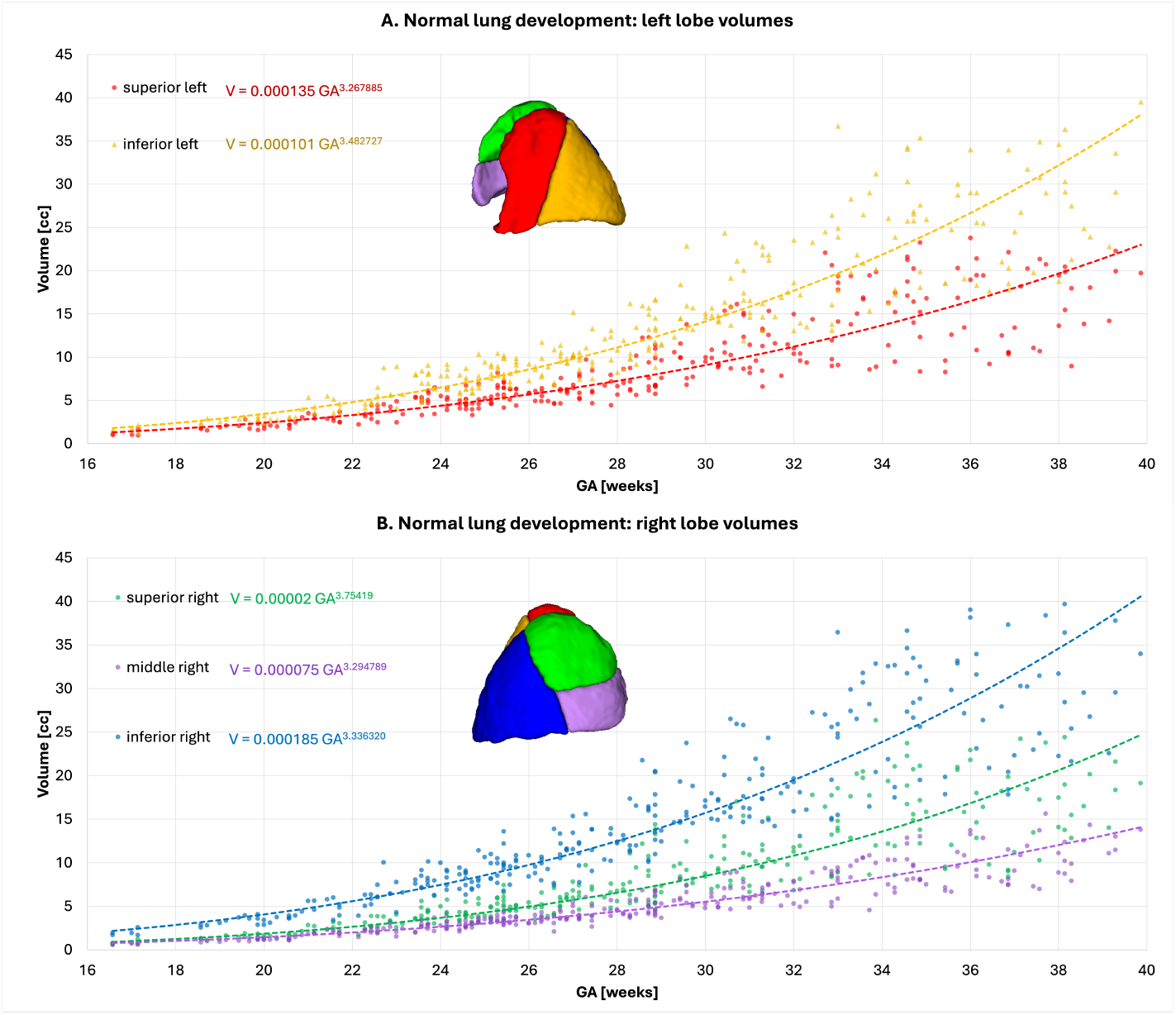
Growth charts created from 155 3T and 135 0.55T normal fetal control 3D T2w datasets for 16 to 39 week GA range: left lung lobes (A); right lung lobes (B).

## 4 Discussion and Conclusions

This work introduced the first solution for automated segmentation of lung lobes for motion-corrected 3D T2w fetal body MRI at various field strengths for both normal and abnormal anatomy. At first, the lobe parcellation protocol was defined in the population-averaged 3D fetal body MRI atlas space. Since the fissures are not normally clearly visible on MRI, the lobe boundaries were delineated based on the expected location vs. supplying vessels and bronchi and the adult CT segmentation protocol as a guidance rather than signal intensity. We used the classical 3D Attention UNet network to segment 5 standard lung lobes. It was trained on 3D fetal body images with manually refined labels propagated from the atlas. The training was based and 104 0.55-3T normal anatomy datasets and a multi-centre 45 CDH datasets with various acquisition protocols. The results of testing on 18 cases are promising and confirm the general feasibility of multi-regional deep learning segmentation for both normal and abnormal CDH lung anatomy at different field strengths. However, training on a wider range of abnormal anatomy cases and potentially more advanced deep learning approach are required for more robust segmentation of ipsilateral lung and other lung anomalies (e.g., congenital pulmonary airway malformation).

This is the first step toward automated multi-regional lung analysis for structural MRI. However, in terms of limitations and future work, a more thorough strategy is required for definition of the lobe boundaries and exclusion of vessels, fluid in pleura as well as investigation of the impact of such factors as GA, lung tissue contrast, SNR, artifacts, thorax orientation, etc. Application of deep learning image restoration and harmonisation should also improve performance on suboptimal image quality cases and different acquisition parameters. The normative charts generated from 290 subjects showed the expected proportions between the left/right lungs, lobe volumes and growth trends. Yet, similarly to the previous works, the variability in values at later GA emphasises the need for deeper analysis of subject-specific factors (parental characteristics, ethnicity, sex, etc.), optimisation of volume normalisation approaches (e.g., total fetal volume [3]) as well individual growth trends in longitudinal datasets.

## Data Availability

All data produced in the present study are available upon reasonable request to the authors.

## Acknowledgments

We thank everyone who was involved in acquisition and analysis of the datasets and all participating mothers and families.

This work was supported by NIHR Advanced Fellowship awarded to Lisa Story [NIHR30166], MRC grant [MR/W019469/1], the Wellcome Trust, Sir Henry Wellcome Fellowship to Jana Hutter [201374/Z/16/Z], DFG Heisenberg funding [502024488], the UKRI, FLF to Jana Hutter [MR/T018119/1], DFG Heisenberg [502024488] the High Tech Agenda Bavaria to J.H., the Wellcome/EPSRC Centre for Medical Engineering at King’s College London [WT 203148/Z/16/Z], the NIHR Clinical Research Facility (CRF) at Guy’s and St Thomas’ and by the National Institute for Health Research Biomedical Research Centre based at Guy’s and St Thomas’ NHS Foundation Trust and King’s College London.

The views expressed are those of the authors and not necessarily those of the NHS, the NIHR or the Department of Health.

## Footnotes

11 SVRTK fetal MRI docker: https://hub.docker.com/r/fetalsvrtk/svrtk

12 ITK-SNAP tool: http://www.itksnap.org/

13 3D fetal body MRI atlas repository: https://gin.g-node.org/kcl_cdb/fetal_body_mri_atlas

14 MIRTK toolbox: https://github.com/BioMedIA/MIRTK

15 Automated SVRTK toolbox: https://github.com/SVRTK/auto-proc-svrtk

16 3D fetal body MRI atlas repository: https://gin.g-node.org/kcl_cdb/fetal_body_mri_atlas

## Notes

### Competing Interest Statement

The authors have declared no competing interest.

### Author Declarations

This study employed fetal MRI data acquired at two different sites: at St.Thomas Hospital, London, UK under the Individualised risk prediction of adverse neonatal outcome in pregnancies that deliver preterm using advanced MRI techniques and machine learning study (REC 21/SS/0082), NANO (REC 22/YH/0210), MEERKAT (REC 21/LO/0742), Placenta Imaging Project (REC 16/LO/1573), Quantification of fetal brain growth and development using MRI (REC 07/H0707/105), IFIND (REC 14/LO/1806) studies and at University Hospitals Leuven, Belgium (Ethics number: S5678). All experiments were performed in accordance with relevant guidelines and regulations. Informed written consent was obtained from all participants. The research ethics committee approval was given by the Health Research Authority boards of: London (including -Fulham, -South East, -Riverside, Dulwich, -West London and GTAC, and -Brent) and, South East Scotland.

## References

1. Aertsen, M., et al.: Fetal mri for dummies: what the fetal medicine specialist should know about acquisitions and sequences. Prenatal Diagnosis 40, 6–17 (2020)

2. Amodeo, I., et al.: The role of mri in the diagnosis and prognostic evaluation of fetuses with congenital diaphragmatic hernia (9 2022)

3. Cannie, M.M., et al.: Fetal body volume at mr imaging to quantify total fetal lung volume: Normal ranges. Radiology 247, 197–203 (4 2008)

4. Cardoso, M.J., et al.: Monai: An open-source framework for deep learning in health-care. arXiv preprint arXiv:2211.02701 (2022)

5. Conte, L., et al.: Congenital diaphragmatic hernia: automatic lung and liver mri segmentation with nnu-net, reproducibility of pyradiomics features, and a machine learning application for the classification of liver herniation. European Journal of Pediatrics 183, 2285–2300 (5 2024)

6. Davidson, J., et al.: Fetal body mri and its application to fetal and neonatal treatment: an illustrative review. The Lancet Child Adol. Health 5, 447–458 (2021)

7. Davidson, J., et al.: Motion corrected fetal body mri provides reliable 3d lung volumes in normal and abnormal fetuses. Prenatal Diagnosis pp. 628–635 (5 2022)

8. Fujii, S., et al.: The bronchial tree of the human embryo: an analysis of variations in the bronchial segments. Journal of Anatomy 237, 311–322 (8 2020)

9. Jani, J., et al.: Value of prenatal mri in the prediction of postnatal outcome in fetuses with diaphragmatic hernia. UOG 32, 793–799 (11 2008)

10. Lassen, B., et al.: Automatic segmentation of the pulmonary lobes from chest ct scans based on fissures, vessels, and bronchi. IEEE TMI 32, 210–222 (2013)

11. Meyers, M.L., et al.: Fetal lung volumes by mri: Normal weekly values from 18 through 38 weeks’ gestation. Am. J. of Roentgenology 211, 432–438 (6 2018)

12. Oktay, O., et al.: Attention u-net: Learning where to look for the pancreas. In: MIDDL 2016 (2018)

13. Payette, K., et al.: An automated pipeline for quantitative t2* fetal body mri and segmentation at low field. In: MICCAI (2023)

14. Perrone, E.E., et al.: Prenatal assessment of congenital diaphragmatic hernia at north american fetal therapy network centers: A continued plea for standardization. Prenatal Diagnosis 41, 200–206 (1 2021)

15. Prayer, F., et al.: Fetal mri radiomics: non-invasive and reproducible quantification of human lung maturity. European Radiology 33, 4205–4213 (2023)

16. Royston, P., Wright, E.: How to construct ‘normal ranges’ for fetal variables. Ultrasound in Obstetrics Gynecology 11, 30–38 (1998)

17. Rypens, F., et al.: Fetal lung volume: Estimation at mr imaging-initial results. Radiology 219, 236–41 (2001)

18. Schittny, J.C.: Development of the lung. Cell and Tissue R. 367, 427 (3 2017)

19. Story, L., et al.: Foetal lung volumes in pregnant women who deliver very preterm: a pilot study. Pediatric Research 87, 1066–1071 (2020)

20. Uus, A., et al.: Deformable slice-to-volume registration for motion correction of fetal body and placenta mri. IEEE TMI 39, 2750–2759

21. Uus, A.U., et al.: Automated 3d reconstruction of the fetal thorax in the standard atlas space from motion-corrupted mri stacks for 21–36 weeks ga range. MedIAn 80 (8 2022)

22. Uus, A.U., et al.: Retrospective motion correction in foetal mri for clinical applications: existing methods, applications and integration into clinical practice. The British Journal of Radiology (8 2022)

23. Uus, A.U., et al.: Automated body organ segmentation, volumetry and population-averaged atlas for 3d motion-corrected t2-weighted fetal body mri. Scientific Reports 14, 6637 (2024)

24. Vimala, V., et al.: Morphological study of fissure and lobes in fetal lungs. International Journal of Anatomy and Research 6, 4959–4962 (2 2018)

25. Wasserthal, J., et al.: Totalsegmentator: Robust segmentation of 104 anatomic structures in ct images. Radiology: Artificial Intelligence 5, e230024 (7 2023)

26. Whitby, E., Gaunt, T.: Fetal lung mri and features predicting post-natal outcome: a scoping review of the current literature (2023)

27. Wilson, L., Whitby, E.H.: Mri prediction of fetal lung volumes and the impact on counselling. Clinical Radiology 78, 955–959 (12 2023)

